# A physically plausible incidence rate for compartmental epidemiological models

**DOI:** 10.1101/2021.06.19.21258779

**Authors:** Thomas Pitschel

**Affiliations:** Goethe University Frankfurt

**Keywords:** compartmental model, SIR-model, non-linear incidence rate function, non-linear force of infection

## Abstract

Motivated by the recent trajectory of SARS-Cov-2 new infection incidences in Germany and other European countries, this note reconsiders the need to use a *non-linear* incidence rate function in deterministic compartmental models for current SARS-Cov-2 epidemic modelling. Employing a homogenous contact model, it derives such function systematically using stochastic arguments. The presented result, which is relevant to modelling of proliferation of arbitrary infectious diseases, integrates well with previous analyses, in particular closes an analytical “gap” mentioned in London and Yorke (1973) and complements the stability related work on incidence rate functions of the form *βI*^*p*^*S*^*q*^ seen for example in Liu, Hethcote and Levin (1987).

## 1 Introduction

The massive vaccination campaigns currently underway in the European countries, and nearly completed in some non-European countries such as Canada, the Unites States, United Kingdom and Israel, have coincided with a rapid decline in new SARS-Cov-2-infection incidences in those countries which is remarkable both in degree and consistency. Given the virus- and contact specific parameter estimates of epidemic models derived just a few months ago, and given the vaccine effectiveness parameters, one may wonder how the currently observed incidence decline could possibly be explained with previously used models.

In Germany, based on data from [1]-[5], the expected overall reduction in infection probability achieved with the current state of vaccination is by a factor of about 0.6391866.^2^ Given that the “unconstrained” (i.e. preventive measures-free) *R*_0_ was estimated at about 3, and the infection probability enters linearly into *R*_0_ in the proliferation models employing a bilinear incidence rate function ^4^, it is unreasonable that the current vaccination state should suffice to effect an incidence decline robust even under lifting of strong preventive measures such as contact restrictions and mask-wearing.

It is therefore hypothesized that the better-than-predicted effect of vaccination is due to another mechanism. The mechanism here supposed is that single-shot (“1s”) vaccinations *raise the tolerance level* for virus exposure in these 1s-vaccinated susceptibles with the effect that *multiple* exposures with virus particles would be required to trigger an infection. (One may regard as suitable criterion the total virus dose an individual is being exposed to over one day, say, and a threshold on this value.) This in turn is deemed to have the consequence that instead of mere reduction of infection probability, an infection is assuredly prevented in specific contact instances in which virus density remains below a certain threshold level (“threshold effect”). As will be seen in the proposed quantitative analysis, the mathematical description of this mechanism leads to a non-linear incidence function in the on-average (=“deterministic”) epidemic modelling. The resulting trajectories then display the larger-than-predicted decline in incidences. As a secondary cause, besides the direct effect of vaccination, the consequential shift of the majority of proliferation events into younger population segments can be stated, which has the same effect of raising the average tolerance level.

The here suggested mechanism has been mentioned as possibly relevant in epidemic modelling, and analytically (nearly) been provisioned for, in much earlier work: In [5], the accounting for multiple exposures is named as a reason to use non-linear incidence functions, and consequently incidence rate functions of form ∼ *I*^*p*^*S*^*q*^ are examined. In [6] the saturation effect, namely that the new infections cannot scale unboundedly linearly with existing number of infectives, is stated to motivate analysis involving non-linear incidence functions, concretely of form *g*(*I*) · *S*, with non-negative bounded differentiable *g* with additional conditions. Most of this previous work however does not focus on a precise translation from physical mechanism of the transmission to a corresponding appropriate mathematical representation of the incidence rate, but often appears to use functional forms guided by tractability or data-matching. What is new in this present note is to give a *systematic* derivation of the non-linearity based on stochastic arguments which properly account for both the saturation and threshold effect. The arguments assume a stylized (in particular: homogenous) contact setting which can be realistic for a substantial part of the contacts occuring in an urbanly situated population (*N* ≈ 100000…1000000). ^5^ Focus is on the overall effect in quantitative terms *under assumption* that the stated mechanism is present.

## 2 Framework epidemic model and derivation of non-linearity *g*

The result to be derived below is supposed to be a building block in the well-known compartmental epidemic models describing the proliferation in terms of the number of susceptible (S), of “exposed”^6^ (E), infectious (I) and removed (R) individuals, for example the one stated as eqn. (2.2) in [11]:

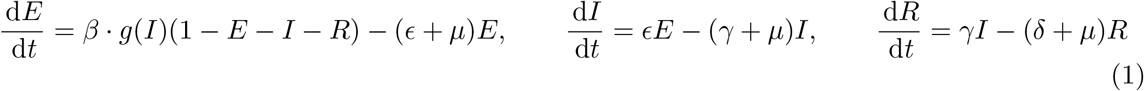

with *S*(*t*) + *E*(*t*) + *I*(*t*) + *R*(*t*) = 1. The time-dependent quantities are understood as *shares* of the respective compartment size relative to the (constant) total population size, and are deemed expected values. The *β* above is a transmission probability and contact mechanism related coefficient, but for the analysis below it will be assumed equal to one. With *S*(*t*) being the share of susceptibles, the *g*(*I*) · *S*(*t*) then takes the meaning of the number of new infections triggered by all infectious among the total population in a unit time interval, divided by *N*.

Historically, the analysis began with the incidence function *g* chosen as linear in *I*, based on the idea that infections caused by an infective are occuring stochastically independently of the other infections (which is approx. true as long as the infectives density is low) and a single contact suffices for infection. At low infectives counts, i.e. *I* ≪ 1, the other members of the population encountered by a specific infective will be nearly disjoint from the ones encountered by any other infective. (I.e. there is no coverage overlap.) Then E(# {“exposed” individuals by any infective)} ≈ 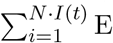 (#{“exposed” individuals by infective *i*}) = *I*(*t*) · const. For larger *I* however, the left-hand side expectation must be bounded by *N*, and the linearity in *I* disappears.

To derive *g*(*I*) systematically for any value of *I*, regard the observation time interval be divided into multiple time slots, and allow any infective to make multiple contacts over the observation interval to other members of the population. Concretely, let *N* · *I*(*t*) be the number of infectives being active at the beginning *t* of the observation time interval, and let each of them make on average *d* contacts with other members during that interval. The contacts are deemed to uniformly randomly covering the population of *N* members, one at a time slot. This is analogous to dialing out a deck of *d* · *N* · *I* cards onto *N* places, for each card choosing one of the places according to the uniform distribution, such that each place may receive multiple cards. We want to compute the expectation of the number of places *K*_*T*_ = *K*_*T*_ (*d* · *N* · *I, N, κ*) among the *N* potential receivers that receive equal or more than *κ* ∈ ℕ cards, corresponding to the number of individuals exposed to an infective at least *κ* times during the observation interval, which is to constitute the condition upon which infection is triggered. (It is assumed here that any contact constitutes an exposure in the sense of contributing to the triggering of an infection; any transmission probability that may exist to describe events of non-exposure inspite of contact is assumed to be absorbed into the value of *d*. Part of the original numerical value of *β* is to appear here.) Since the susceptibles are in turn independently distributed among the members of the population, it then follows the expected number of infections triggered in the interval is E(*K*_*I*_) = E(*K*_*T*_) · *S*(*t*).

The probability *p*_0_ of a specific member of the population being “hit” by zero infectives is given by ((*N* − 1) / *N*) ^*dNI*^ We will write *d* · *I* = *Ĩ* and focus on large *N*. Then *p*_0_ ≈ exp(−*Ĩ*). The probability *p*_1_ of a specific member being hit once by an infective is

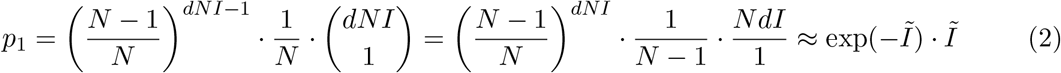

Generally, the probability *p*_*k*_ of a population member being exposed exactly *k* times is found to be

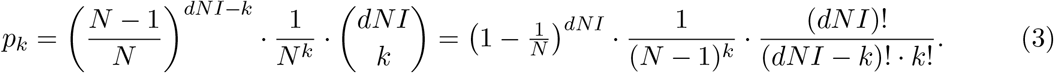

For *k* small compared to *N* and compared to *Ĩ* · *N*, it is 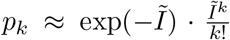. (The random variable counting the exposures at a specific population member approximately is found to be Poisson distributed with intensity *Ĩ*.) The approximate values are normalization-consistent. The probability of a specific member being exposed at least once consequently is (1 − *p*_0_) ≈ 1 −exp(−*Ĩ*); of being exposed at least *κ* times it is 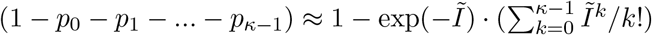.

Based on these elementary arguments, one derives the expected values as follows: Since all members of the population are here deemed equal in their role as potential virus receivers, it is

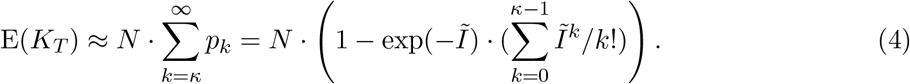

As argued before, E(*K*_*I*_) = E(*K*_*T*_) · *S*(*t*). The incidence function *g* derived by above arguments thus is

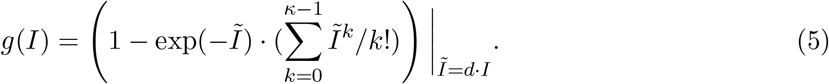

### 2.1 Discussion

As intended, the incidence rate function here is bounded by 1 even as *d* · *I* increases to infinity. Further, the historical bilinear form *βIS* is recovered as special case by observing that for *κ* = 1, i.e. a single exposure is deemed sufficient for triggering an infection, the first order approximation of the exponential at *Ĩ* small yields

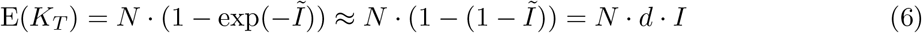

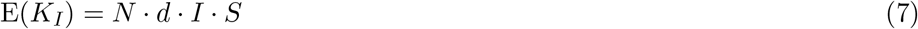

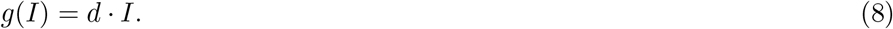

The next higher order approximation can be verified to equal E(*K*_*T*_) = *N* · (*Ĩ* − *Ĩ*^2^/2 = *N* · *d* · *I*(1 − *dI*/2), yielding the first component accounting for coverage overlap leading to the saturation phenomenon. The stochastic analysis of the here employed homogenous contact mechanism is found to motivate well the non-linear modification of the historical form introduced in [12] (pg 471) based on empirical grounds (“to eliminate systematic differences [to observed data]”), with additional benefit of attaching a meaning to the arising coefficient.

#### *The case of multiple required exposures (κ* > 1)

It is easy to see that the leading monomial in *g*, for small values of *Ĩ*, will always have degree *κ*. Exemplarily, one has as approximations for small *Ĩ*:

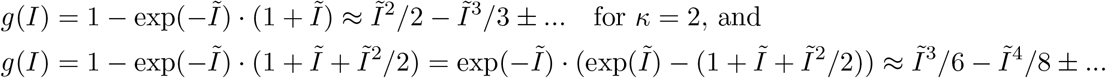

for *κ* = 3. This leads, at small *Ĩ*, to a polynomial behaviour of the incidence function, namely *g*(*I*)· *S* ∼ *I*^*κ*^ · *S*, in line with the form previously analysed in [5] and subsequent work. Based on the host response characteristics determined for COVID-19 in medical literature and shortly described in footnote 5, one concludes that for COVID-19 such super-linear incidence rate behaviour will be relevant rather in younger population segments. These are exactly the segments that constitute the currently remaining “theatre of proliferation” after the advancement of the vaccination campaigns.

## Data Availability

Only already publicly available data was used.

https://www.rki.de/DE/Content/InfAZ/N/Neuartiges_Coronavirus/Daten/Impfquotenmonitoring.html

https://impfdashboard.de/

## Footnotes

2 In Germany, according to [1], as of 29th May 2021 there had been administered about 45.3 mio anti-SARS-Cov-2 vaccine doses, and about 14.20 mio persons (17.1% of the population) had been completely vaccinated. In about 42.2% of the population at least one vaccine shot had been administered. According to [2], about 50.5 mio doses had been delivered until 23th May, and of those 97.5% had been administered until 28th May; The distribution of vaccine types among those delivered doses are [2]: 70.5% of BioNTech/Pfizer (BNT162b2), 19.2% of AstraZeneca (AZD1222), 9.4% of Moderna (mRNA-1273), 0.9% of Johnson&Johnson (JNJ-78436735). Except from the JNJ-78436735, which requires one shot to be administered to completion, the three other vaccines require two shots. The reported effectiveness of the vaccines in preventing (symptomatic^3^) infection was stated as: 82%/95% after first/second shot of BNT162b2 [3], and 76%/81.3% after first/second shot of AZD1222 [4]. For Moderna’s mRNA-1273 we assume the same effectiveness numbers as for BNT162b2. If we regard the effectiveness denoting an equivalent reduction of *probability* of infection, and assume vaccine type distribution in the administered doses equals those in the delivered doses, and assume equal vaccination progress in each vaccine type, and neglect J&J vaccines from the calculation, then one must tend to conclude that the overall reduction of infection probability achieved by the current state of vaccination, in Germany, is by an aggregate factor of 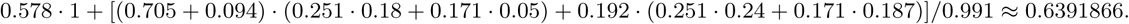 Here, the 0.578 represents the share of the not-yet-vaccinated population in which no reduction takes place. The 0.991 adjusts for Johnson&Johnson numbers being omitted.

4 Have *R*_0_ = *µ · T*_*I*_ = *d · p · T*_*I*_, where *µ* is a transmission coefficient per time unit, *T*_*I*_ is the average infectiousness period, *d* is a contact degree and *p* is an aggregate transmission and reception probability.

5 In the author’s opinion the arguments stated in [7] for focussing merely on concave incidence rates (on biological grounds) are not plausible: The threshold effect will be relevant for any communicable disease in which the counts of transmitted disease carriers can be reduced, say by physical preventive measures (e.g. using masks), or the tolerance level can be raised, as long as a dose-response relationship is at work which has sufficient smoothness. This dose-dependency of the response (and ultimately: of secondary infectiousness) is present e.g. for COVID-19 at least in the younger (i.e. usually more healthy) population ([8, 9] with [10] (pg 1043)).

6 As will become clear later, this group in the present text would rather be called “the individuals in which infection is triggered but who are not yet infectious”.

